# Background noise in an Emergency Department: an observational study from staff and patient perspectives

**DOI:** 10.1101/2022.05.20.22275148

**Authors:** Murad Emar, Ella Smith, Timothy J Coats

**Affiliations:** Emergency Department, University Hospitals of Leicester NHS Trust, Leicester, UK; University of Bristol, UK; Emergency Medicine, Dept of Cardiovascular Sciences, University of Leicester, Leicester, UK

**Author notes:** Corresponding author: Prof T Coats, Emergency Medicine, Dept of Cardiovascular Science, University of Leicester, University Road, Leicester, UK, LE1 7RH. Sources of funding: None.

**Keywords:** Emergency Medicine, Noise, Emergency Service (Hospital), Communication

## Abstract

**Background and importance:** Noise is a contributing factor to mis-communication, poor sleep patterns and stress in healthcare. There has been little research on noise in the Emergency Department (ED).

**Objective(s):** (1) To identify the noise levels experienced by staff and patients in different areas of an emergency department over the 24 hour cycle, (2) to examine the impact of cubicle doors on the background noise experienced by the patient, and (3) to assess the impact of monitor alarms on staff and patient noise levels.

**Design:** Observational study.

**Setting:** A large urban teaching hospital Emergency Department.

**Measures and analysis:** Using a standard protocol monitoring of staff and patient experience of noise was carried out in 3 areas of the ED (a resuscitation room, an area of patient cubicles with solid doors and and an area of patient cubicles with curtains).

The overall distributions of noise levels in each area were described and circadian variation plotted. The proportion of time that background noise was above key cutoff values known to impair communication was calculated (45dB and 65dB).

Non-parametric methods were used to compare: (1) a patient cubicle with curtains compared to a solid door, (2) having the door open or closed, and (3) staff and patient exposure a monitor alarm.

**Main results:** Noise was greater than 45dB for staff between 76% and 96% of the time (30% to 100% for patients). There was little difference across the 24hr cycle. A door decreased the noise experienced by patients, but only if left closed. In the resuscitation rooms monitor alarms were much louder for patients than for staff.

**Conclusion:** Noise levels likely to impair communication are present in the ED for most of the time. Staff awareness and improved design of both buildings and equipment might mitigate this negative acoustic environment.

## Introduction

There is little published data about background noise in ED, with the existing evidence using a heterogenous group of methodologies which makes interpretation difficult. There is no published comparison of the staff and patient acoustic environments.

Sound is a wave motion carrying energy through a medium (such as air) which then exerts force (pressure) on a surface. The human ear can hear a wide range of sound pressure (from 0.00002 pascals up to 200 pascals) so for convenience a logarithmic scale is normally used - expressing the sound pressure level as decibels (dB). On this logarithmic scale (Table 1) for every 10 decibels of increase, loudness is 10 times greater (a 3dB increase is roughly doubling the loudness). Therefore, a small difference in sound pressure level (dB) corresponds to a much larger difference in perceived loudness(1).

**Table 1.** Examples of noise levels in decibels (dB) - a logarithmic scale so each group is 10 times the loudness.

|  |  |
| --- | --- |
| 30 dB | Voice whispering |
| 40 dB | Library |
| 50 dB | Normal speech at 1m |
| 60 dB | Loud speech at 1m |
| 70 dB | Loud television |
| 80 dB | Kitchen mixer, Alarm clock, Voice shouting |
| 90 dB | Workman drilling, Loudest shout |
| 100 dB | Fighter jet at 300m, Chainsaw |

The World Health Organisation Community Noise Guidelines(2) suggest that for complex communication the voice should be at least 15 dB louder than the background, so at 1m the loud speech level of 60dB is 100% intelligible in background noise levels of up to 45 dB (shouting will be intelligible up to a background noise level of 65dB). The guidance suggests that hospital rooms should have a background noise level below 35dB (30dB at night).

Most studies of background noise in hospitals(3-6) have concentrated on patient impacts (sleep patterns, wellbeing and satisfaction) rather than impact on staff communication. A large variety of methods of measurement have been used, which makes comparison of results difficult(7).

However, high noise levels with negative associations (stress, sleep deprivation, cardiovascular effects, and anxiety) have been found in the hospital ward(8), maternity unit(9), critical care unit(10), neonatal intensive care, operating theatre(11), and paediatric clinic(12).

Previous measurements of noise in ED(13-15) have demonstrated noise levels (50 to 70 dB) high enough to raise concern about communication interference and subsequent disruption of complex procedures(16, 17), teaching(18) and decision-making (19-20). The highest noise levels (a medical helicopter landing or use of an orthopaedic cast saw) were above recommended occupational exposure levels(21) suggesting that protective equipment should be used. This previous work has led to recommendations about strategies to improve the acoustic environment in emergency care(15, 22) and to improve sleep in patients ‘boarding’ overnight in ED while waiting for a hospital bed(23). Previous work had highlighted the contribution of monitor alarms to noise in hospitals(24), but had not compared the effect on patients and staff.

The current literature and recommendations are based on relatively few studies of noise in the Emergency Department with the use of non-standard methods making it difficult to derive conclusions(7) and with none comparing different areas of the ED, comparing the noise experience of staff and patients or describing the effect of patient cubicle design. This study therefore aimed (a) To identify the noise levels experienced by staff and patients in different areas of an emergency department over the 24 hour cycle, (b) to examine the impact of cubicle doors on the background noise experienced by the patient, and (c) to assess the impact of monitor alarms on staff and patient noise levels.

## Methods

The study was undertaken in a single large UK urban teaching hospital (ED attendances 240,000 per year). The measurements were undertaken in the main three areas of the emergency department: (a) the ‘Red majors’ area which has 32 cubicles, each with brick walls and a sliding wall-to-wall glass fronted door system, arranged in a square with staff workstations in the centre of the square, (b) the ‘Blue majors’ area which has 35 cubicles separated by brick walls with a curtain across the front of each cubicle, and one centrally located staff workstation, and (c) the Resuscitation room which has 12 cubicles with brick walls and glass doors with one centrally located staff workstation.

Measurement methods followed the recommendations for sound measurement in hospitals(7). Two sound metre devices were used to carry out the measurements; a Martindale SPL82 and an Amgaze GM1356. Both were calibrated at 94dB twice a week using a Martindale SPC70 sound calibrator. In accordance with standard recommendations a dBA frequency weighting and a ‘fast’ time weighting were used(7). The recording interval in both instruments was set to every 2 seconds. During recordings the microphones were mounted on a tripod and placed near to the patient / staff locations, but away from surfaces such as walls and floors, to eliminate errors caused by reflection and to ensure any noise source was not obstructed.

To identify the overall noise levels experienced by staff and patients, four 24 hour recordings were made in each of the areas of the emergency department - Red majors (cubicles with doors), Blue majors (cubicles with curtains), and the Emergency (Resuscitation) room.

Recordings were made at the staff workstation in each area and inside the nearest patient cubicle. The overall results of 96 hours of recordings in each area were displayed as ‘violin plots’ to show the distribution of background noise measurements, with median and interquartile range calculated. To show the circadian variation the median and IQ range was calculated and plotted for each hour of the day. The data were further described by calculating the proportion of time when the background noise was above a cutoff of 45dB (chosen as with background noise above this level loud speech (60dB) is unlikely to be fully intelligible as the signal to noise gap is <15dB. The proportion of time that the background noise was above 65 dB was also calculated, as above this level loud even shouting (80dB) is unlikely to give a 15 dB signal / noise gap, and so will not be fully intelligible.

To examine the impact of cubicle doors on the background noise experienced by the patient both ‘experimental’ and ‘real life’ recordings were made. For the ‘experimental’ situation the noise level was monitored inside a patient cubicle in sequential sets of 5-minute recordings with the door alternating closed and open. The noise levels were averaged over each 5-minute period and a graph of the resulting sequence noise levels was created. All of the recordings were then grouped into door open or door closed, and the statistical significance of differences was tested using a Kolmogorov-Smirnov test. For the ‘real life’ recording, simultaneous recordings of 2 hours duration were made inside and outside of a cubicle. Two sets of recordings were made, one set recorded in the ‘Red majors’ area (inside and outside of a patient cubicle with a glass door), the other set being recorded in the ‘Blue majors’ area (inside and outside of a patient cubicle with a curtain). Patients were present in the cubicles and no constraint was placed on staff activity during the 2 hour ‘real life’ recordings. The simultaneously recorded paired data (inside and outside of each type of cubicle) was compared using a Wilcoxon matched pairs signed rank test.

To look at the effect of patient monitor alarms on the noise environment experienced by both staff and patient simultaneously recording of noise levels were made both inside an Emergency Room (Resuscitation) cubicle (with the door closed and no patient present) and at the staff workstation in sequential sets of 10-minute recordings with the patient monitor alarm alternating alarm sounding and alarm off (the alarm sound being generated by both the monitor in the patient cubicle and at a central monitoring workstation). The median sound volume in the patient cubicle and at staff desk during each part of this sequence was calculated and plotted, and the difference between ‘alarm’ and ‘no alarm’ noise levels at patient and staff location were compared using a Kolmogorov-Smirnov test.

## Results

The data for the 24hr recordings showed that apart from the patient cubicle in the Emergency (Resuscitation) Room there was a similar skewed distribution of noise in each area (Figure 1).

**Figure 1.**
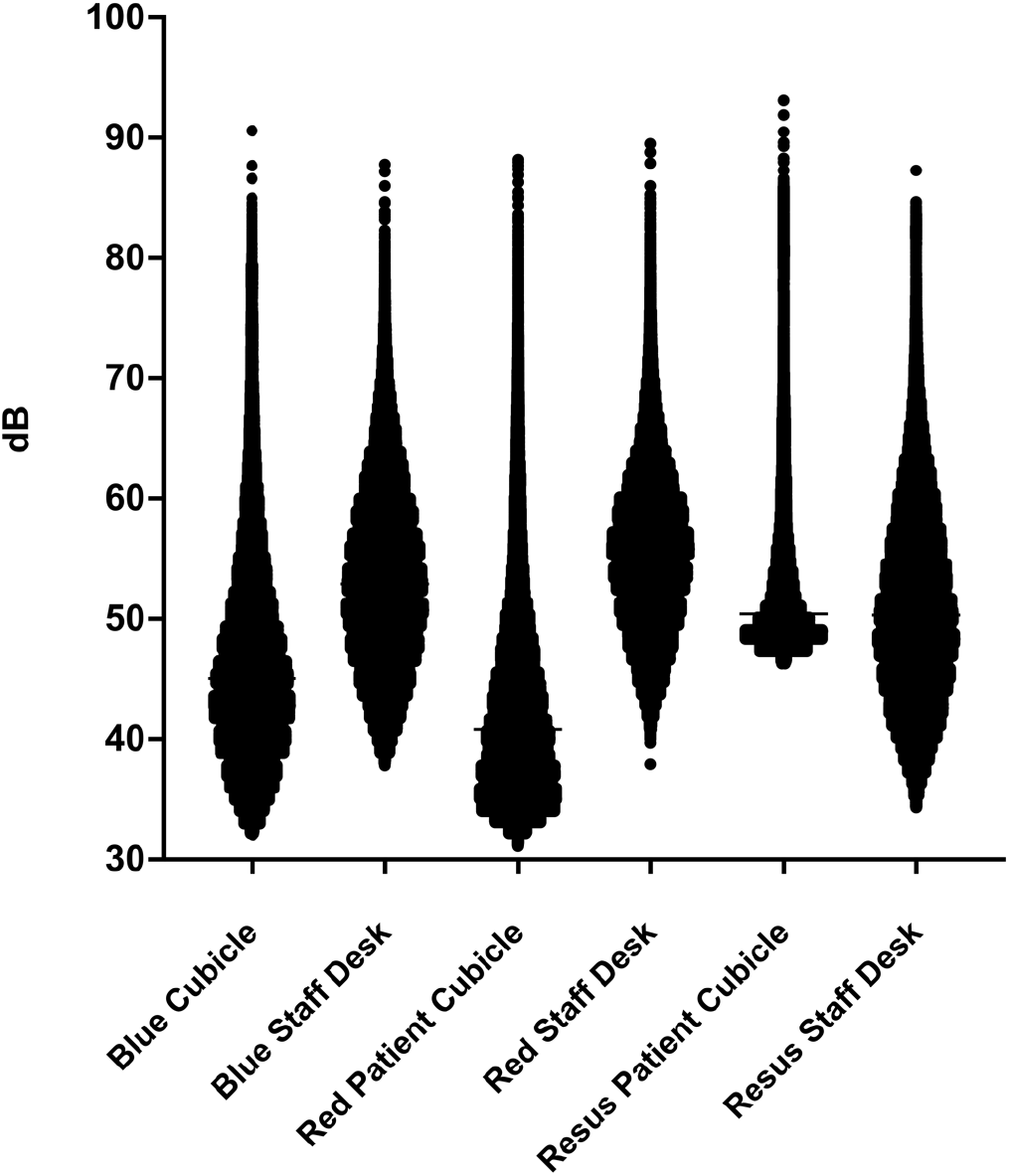
Distribution of noise levels in the ED.

The summary results for each area (Table 2) showed that between 76% and 96% of the time noise levels were >45dB (the level at which staff would have to speak with a loud voice to be fully intelligible). For between 5% and 7% of the time noise was >65dB (when even shouting would not give full intelligibility).

**Table 2.**
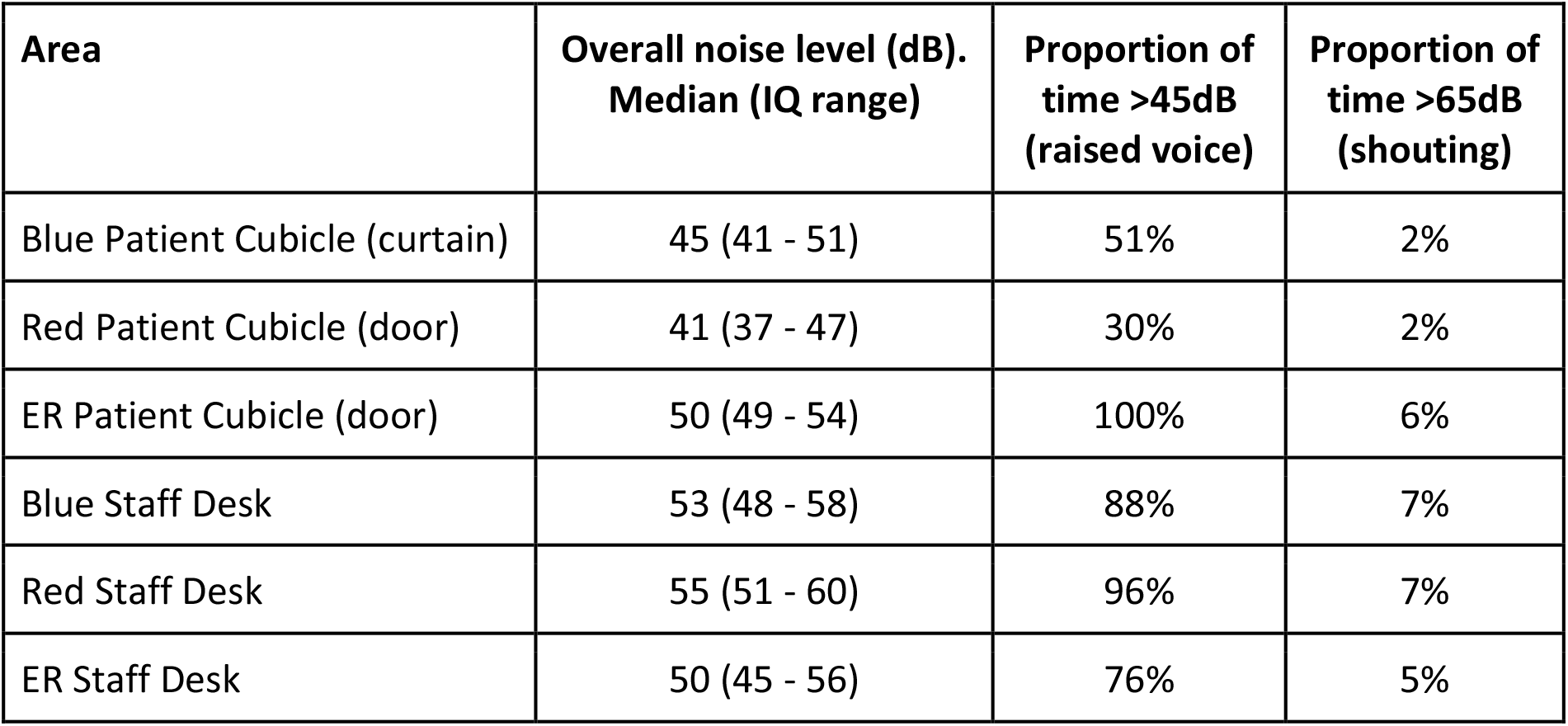
Summary of results.

The volume of background noise averaged over the 4 days of recording in each area was relatively consistent over a 24hr cycle in both staff and patient areas (Figure 2). Staff in all areas were consistently experiencing background noise of 50 to 60 dB. Patients in the Emergency Room had a similar noise exposure to staff, however in the ‘Red majors’ and ‘Blue majors’ the patient’s noise experience was consistently lower than the staff’s, with this difference being greater in ‘Red majors’ (where there are doors rather than curtains on each cubicle).

**Figure 2.**
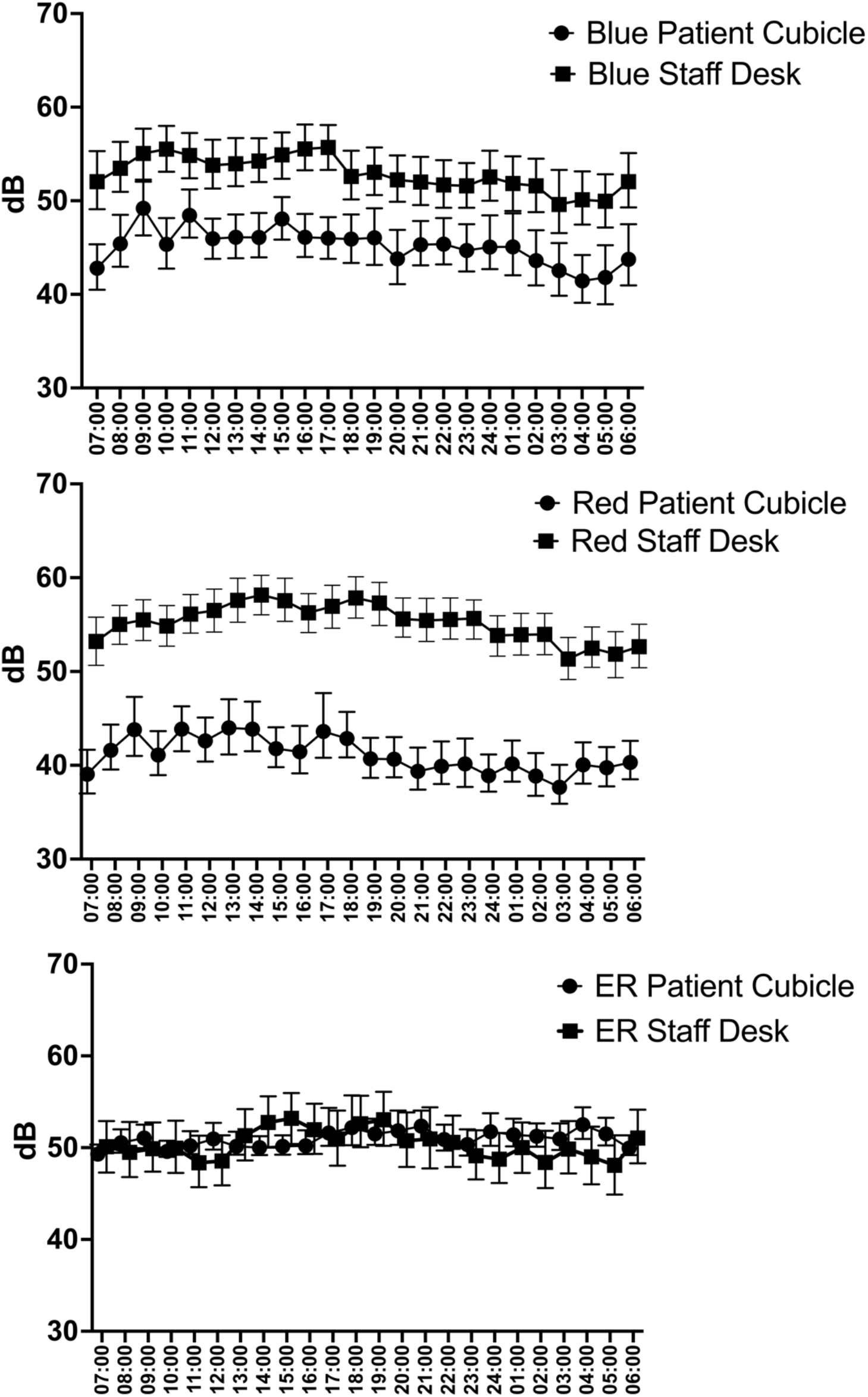
Circadian variation in background noise.

The ‘experimental’ recordings of the impact of cubicle doors on the background noise experienced by the patient showed that for cubicles with a glass door measurement inside of the patient cubicle with opening and closing of the cubicle door gave a clear pattern (Figure 3), The overall median background noise level with the door open was 51dB (IQ range 48 to 54) with the door closed and 44dB (IQ range 42 to 46), the difference between medians being 7dB. This difference was statistically significant (p<0.0001).

**Figure 3.**
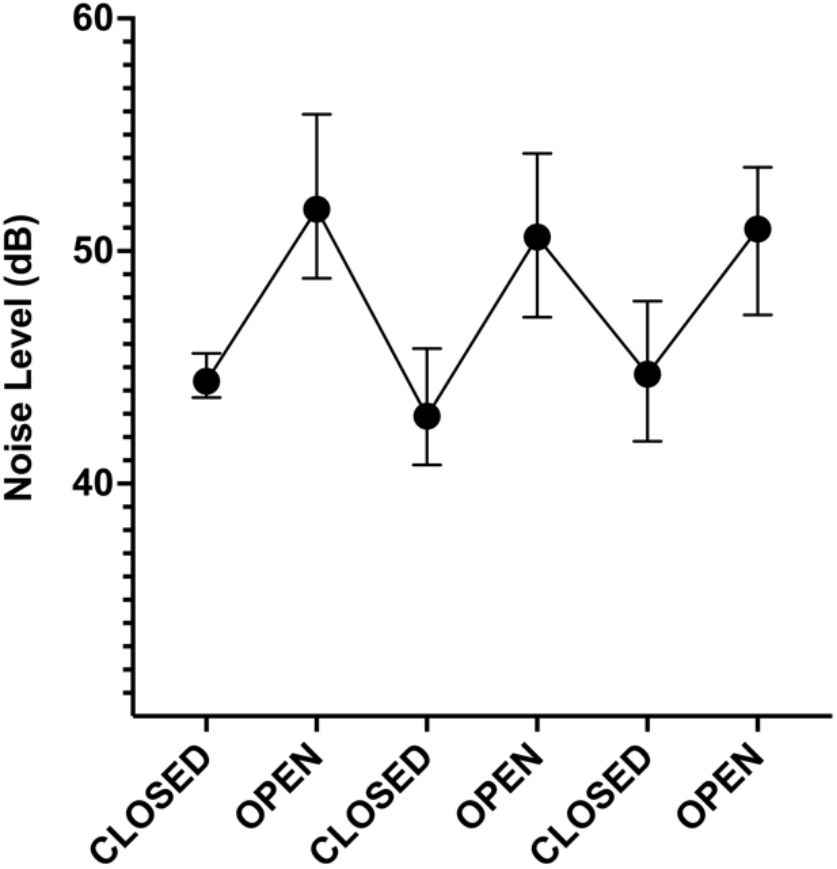
Effect of glass door being open or closed on noise inside a patient cubicle.

The ‘real life’ recordings (Figure 4) of the effect of cubicle doors did not show as large an effect as the ‘experimental’ recordings with a glass door giving a 4dB difference with median reading of 57dB (IQ 53 - 62) outside and 53 (IQ 46 to 60) inside. The noise outside of a curtained cubicle was 54dB (IQ 49 - 59) compared to 50db (IQ 46 - 75) inside the cubicle (also 4dB difference). In real life the inside of a patient cubicle was 4dB quieter whether or not the cubicle had doors (compared with an 7dB decrease in noise when the doors were closed in the ‘experimental’ recordings). During the ‘real life’ recordings the researcher noticed that ED staff rarely completely closed the cubicle doors.

**Figure 4.**
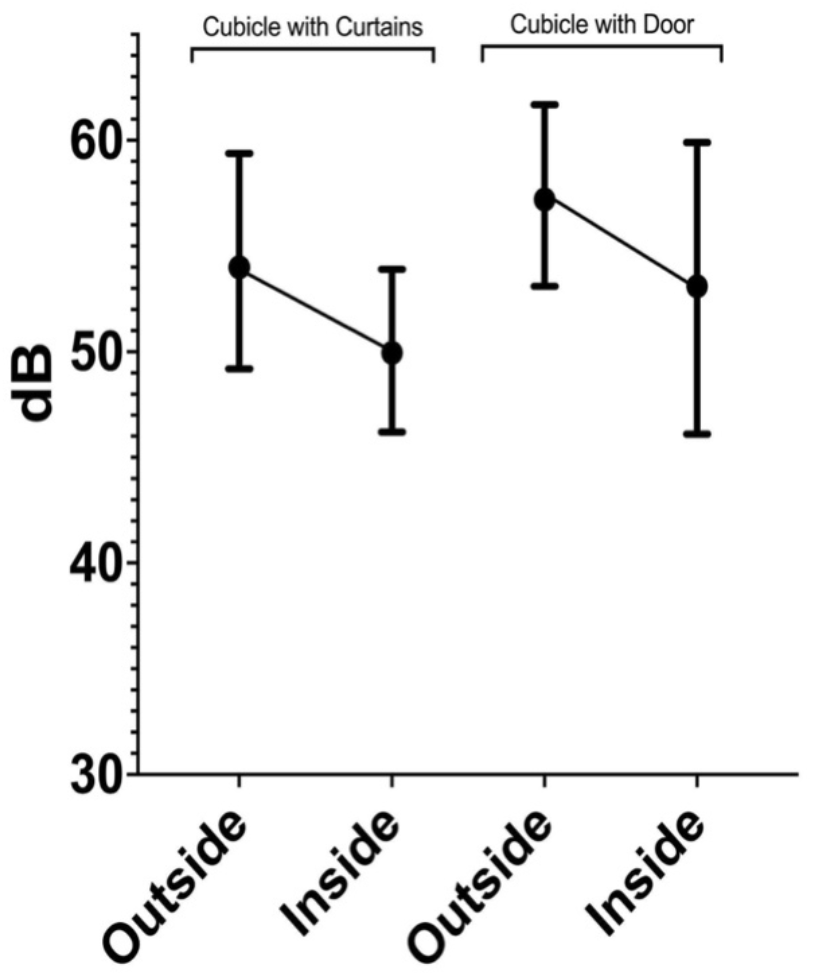
Simultaneous recordings of background noise inside and outside of cubicles with curtain and door.

The simultaneous recording of perception of monitor alarms by staff (recording at staff desk) and patient (recorded in patient cubicle) showed a large difference, with no change in the overall noise level at the staff desk when the alarm activated (50.6dB v 51.2dB, p=ns), yet a very large difference within the patient cubicle (39dB without alarm v 74dB with alarm, p=<0.0001) (Figure 5).

**Figure 5.**
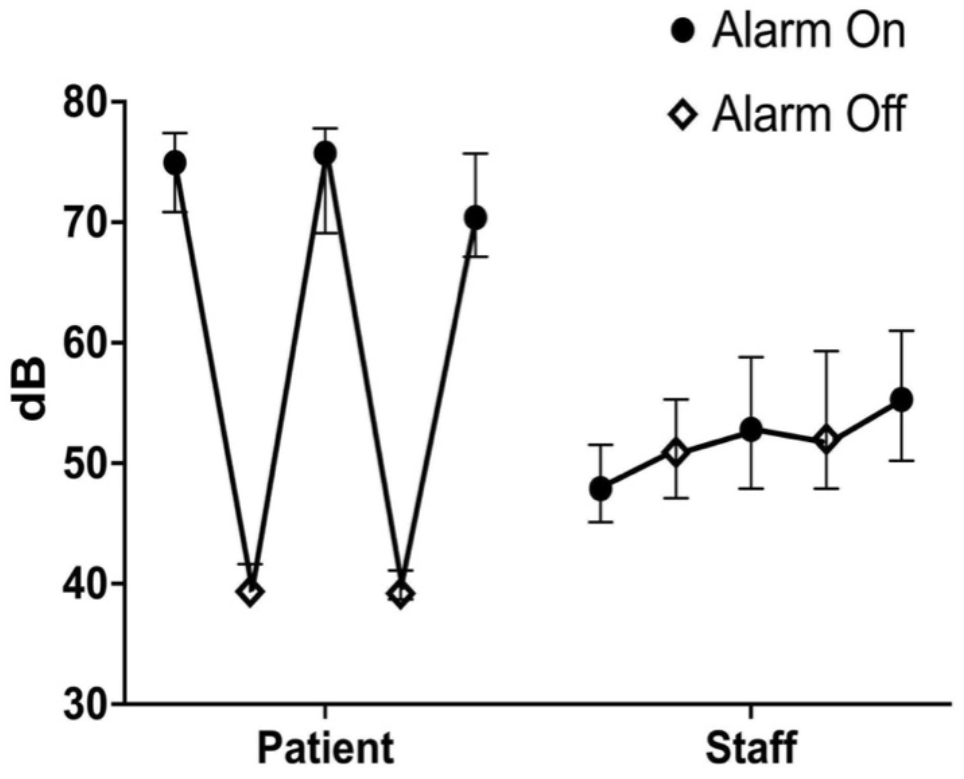
Difference between patient and staff experience of monitor alarm.

## Discussion

This study is the first to examine both staff and patient experience of noise across different areas of an Emergency Department. In all the areas studied levels of background noise were found which are likely to impair communication, decrease patient comfort and lead to staff fatigue. In general, the sources of background noise are outside of the patient’s cubicle, so being further from the noise sources means that patients in general have slightly less (by 4 to 7 dB) exposure to noise than staff. However, this is still well above the recommended 30dB level needed for overnight rest. This may not have mattered in the past when patients moved through the ED onto a hospital ward, however the increasing numbers of patients ‘boarding’ in the ED means that overnight rest is now an important part of ED care, especially to those vulnerable to noise due to confusion, neurodivergence, mental health problems or dementia. It seems that strategies long ago implemented on wards(25) to improve patient’s sleep now need to be implemented in the ED.

The design and use of the ED has an effect on the patient’s experience of noise. A previous study in a neonatal intensive care unit(26) found only a small difference in noise (3 dB) between closed ‘pods’ and open ‘cubicles’. Our results may explain this difference, as the way in which staff use the cubicle doors has an impact - reduction in noise was greater (7dB) during an experimental use of doors than when the doors were used in real life (a 4dB reduction). From observation this difference may be because staff rarely fully closed the doors (anecdotally some patients also do not like the claustrophobic feeling of being in a room with fully closed doors). Doors only protect the patient from noise generated outside of the cubicle, they give no reduction for sounds generated by the patient themselves or coming from equipment within the room (such as equipment alarms).

Alarm fatigue (ignoring frequently sounding alarms which only rarely indicate a serious change in condition) is well known in emergency care and impairs patient safety(27). This study is the first to compare the patient and staff experience of alarms in the ED, showing that the patients experience uncomfortably loud sounds from the equipment inside the cubicle (>70dB), whereas the alarms from the central monitoring console hardly register above the background noise for staff. It is likely that the louder alarm sound for the patient is because they are close to the monitor and located in a confined space with highly reflective hard, flat surfaces (walls and glass doors). In contrast the external central monitor to alert staff is in a large space with fewer reflective surfaces and positioned about 4m from the staff desk. There is the potential to improve patient experience and safety by including acoustic planning in ED design(28).

There was a marked difference in the pattern seen in the Emergency Room compared with the two other areas studied – with both patients and staff experiencing higher levels of noise. From observation this may be because in ‘Red majors’ and ‘Blue majors’ areas most of the noise is generated external to the patient’s cubicle (at or near the staff workstations) by staff actions (conversations and movement) and processes (phone calls, tannoy calls). In contrast, in the Emergency Room the noise that the patient experienced was from inside their room – in addition to the alarms and conversations with staff, we noticed a continuous 40dB -45dB background noise. This originated from the gas scavenging vacuum for the anaesthetic machines, which was always on and could not be individually controlled. Whilst this might not be a universal issue, it was another example of the acoustic environment not being considered in ED design.

The background noise levels found are likely to impair communication, particularly for complex sentences, and particularly if communication is in a second language, heavily accented or there is hearing loss or dementia. Impaired communication has been associated with error – in fact both investigators could easily recall recent anecdotes: In one instance a clinician was breaking news of a death through a telephone call as a noisy trolley passed by, and it was embarrassingly unclear if the relative had actually heard the word “died”. In another instance, a junior doctor requested a chest CT (“CT”) scan rather than CT pulmonary angiogram (“CTPA”), as the alarm from a monitor had made the consultant’s words unclear. There is the potential to improve patient safety by interventions to reduce noise in emergency care, such as better acoustic design(4), writing rather than talking(29), warning of high noise levels(30), or raising staff awareness(15).

Only measuring in one ED was a limitation of this project, but the ED studied does not have unique features in design and function so the results are likely to be generalisable. Staff may have modified their behaviour as they knew that recordings were being made, however the length of the recordings (longer than any previous study) was designed to habituate the staff to the recording devices. Only one cubicle was tested in each area (the nearest to the staff workstation), however there could be significant variation between patient cubicles depending on proximity to potential noise sources. No account was taken of how the sound environment in a particular location may also depend on the patient - for example an unstable patient could experience more noise from frequent alarms. There may also be unmeasured variation from nearby patients - either unusually quiet and withdrawn or aggressive, drunk and shouting.

The ED is known to be a noisy environment. This study has quantified staff and patient experience of noise throughout the 24 hour cycle, which is constant, above recommended levels and likely to give both fatigue and communication error. Doors rather than curtains on patient cubicles seem to have little effect, as in real life the doors are often left partially open. Equipment design (in particular the effect of monitor alarms), the design of the built environment and staff awareness and training seem to be key areas for future research on interventions to decrease the adverse effects of a noisy environment on both patients and staff.

## Supporting information

STROBE Checklist

## Data Availability

All data produced in the present study are available upon reasonable request to the authors.

